# Lossless Distributed Linear Mixed Model with Application to Integration of Heterogeneous Healthcare Data

**DOI:** 10.1101/2020.11.16.20230730

**Authors:** Chongliang Luo, Md. Nazmul Islam, Natalie E. Sheils, Jenna Reps, John Buresh, Rui Duan, Jiayi Tong, Mackenzie Edmondson, Martijn J. Schumie, Yong Chen

## Abstract

Linear mixed models (LMMs) are commonly used in many areas including epidemiology for analyzing multi-site data with heterogeneous site-specific random effects. However, due to the regulation of protecting patients’ privacy, sensitive individual patient data (IPD) are usually not allowed to be shared across sites. In this paper we propose a novel algorithm for distributed linear mixed models (DLMMs). Our proposed DLMM algorithm can achieve exactly the same results as if we had pooled IPD from all sites, hence the lossless property. The DLMM algorithm requires each site to contribute some aggregated data (AD) in only one iteration. We apply the proposed DLMM algorithm to analyze the association of length of stay of COVID-19 hospitalization with demographic and clinical characteristics using the administrative claims database from the UnitedHealth Group Clinical Research Database.

## 1. Introduction

The COVID-19 outbreak has become a pandemic, causing a large increase in mortality and posing a heavy burden to the healthcare system. Much research has been done on treatment efficacy and adverse clinical outcomes [1-5] and much remains to be done. As studies continue to be conducted and published, multi-site collaboration is demanded for evidence synthesis [3,4]. Multi-site studies based on healthcare data, including the electronic health record (EHR) and claims data, can integrate clinical information across multiple sites or systems to improve estimation and predictive performance due to use of a larger and more inclusive sample from the population of interest.

One primary challenge for multi-site collaboration is preserving the privacy of protected health information. Sensitive individual patient data (IPD) including the patient’s identity, diagnoses and treatments are usually not allowed under privacy regulation to be shared across networks. Existing approaches to performing multi-site studies, e.g. distributed algorithms, have the drawback of biased estimation [12] or communication burden due to the requirement of iterative transmission of summarized data [14,15]. Specifically, for ordinary linear regression, identical results with pooled analysis can be obtained by lossless compression [7]. Another important challenge of analyzing multi-site data is the heterogeneity of data distribution across sites. Many existing approaches for multi-site analysis assume the data are homogeneously distributed across sites. This assumption may be violated in practical scenarios and thus make the model vulnerable in estimation and hypothesis testing. For instance, in this manuscript we consider the length of stay of the hospitalization due to COVID-19 as the continuous outcome. Heterogeneity across countries, regions, and sub-populations has been reported in the literature [2]. The aforementioned lossless compression approach [7] for ordinary linear regression, fails to take the heterogeneity into account.

In this paper we propose a novel algorithm for distributed linear mixed models (DLMMs). Linear mixed models (LMMs) are commonly used in many areas including epidemiology for analyzing multi-site data with heterogeneity. The model assumes site-specific random effects of the covariates (and intercept) on a continuous outcome. To the best of our knowledge, there is no existing approach for fitting LMMs in a distributed manner, see Figure 1 for the comparison of several approaches in this context. Our proposed distributed LMM can achieve exactly the same results as if we had pooled individual patient data from all sites, hence the lossless property. These lossless results can be obtained by requiring the sharing of summary statistics from each site in only one iteration. We apply the proposed DLMM to analyze the association of length of stay of COVID-19 hospitalization with demographic and clinical characteristics using the administrative claims database for Medicare Advantage members from a large US Health insurance provider (Appendix Figure 2A).

**Figure 1.**
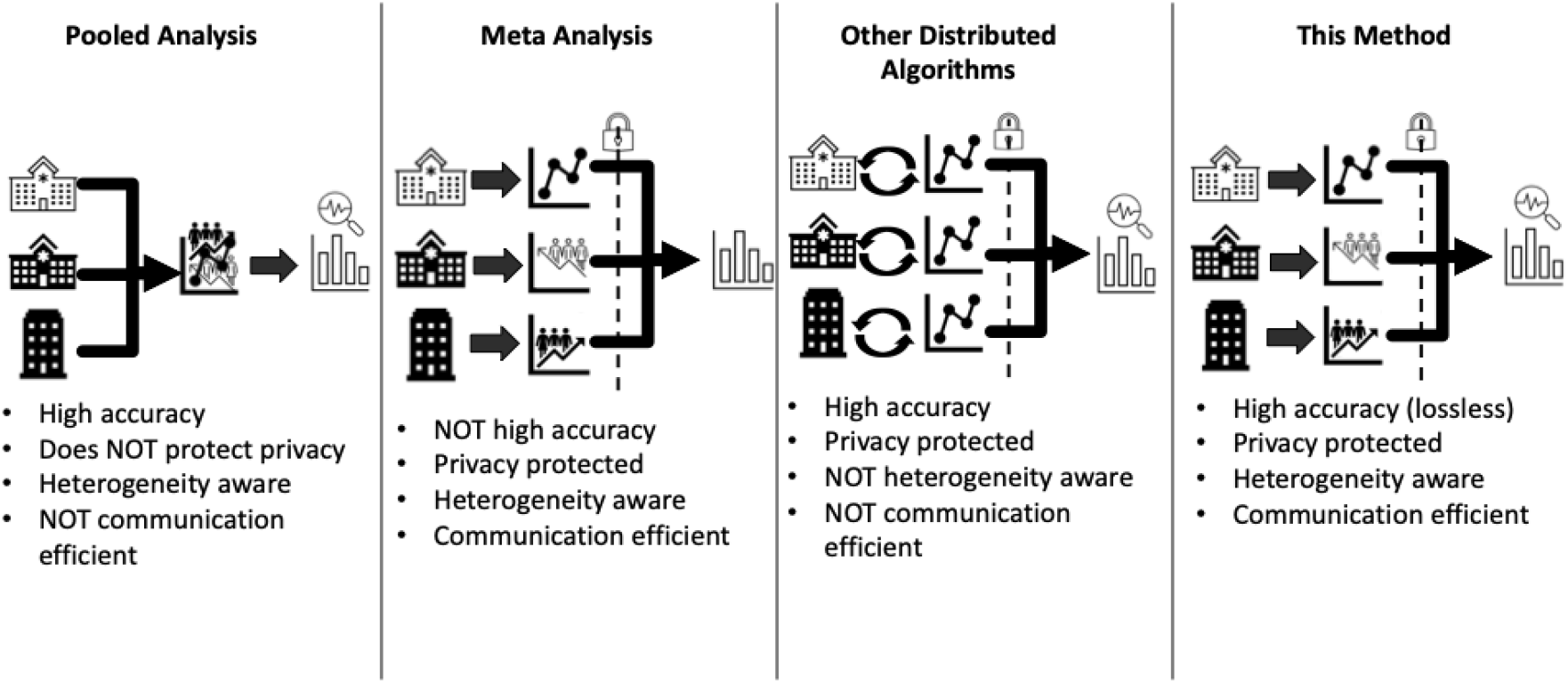
Comparison of several approaches for linear regression analysis of multi-site EHR data with heterogeneity.

## 2. Method

### 2.1 Linear mixed model

Due to the heterogeneity of data across sites, the effects of the covariates on the outcome among sites in the linear regression model may not always be the same [7]. A linear mixed model is thus often used. Assume for the *j*^th^ patient at the *i*^th^ site, *y*_*ij*_ is the continuous outcome, *x*_*i*j_is the *p*-dimensional covariate vector and *β* is the vector of fixed effects, *z*_*i*j_is the *q*-dimensional covariate vector having random effect *u*_*i*_, and *ϵ*_*i*j_is the random error.

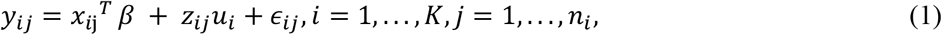

where *u*_*i*_ ∼ *N*(0, *V*), *ϵ*_*ij*_ ∼ *N*(0, σ^2^). The random effects covariates *z*_*i*j_ can be part or all of *x*_*ij*_, or constant if random intercept only. The random effect covariance matrix *V* can admit certain structures with unknown parameters. For instance we can assume the random effects are independent, i.e. *V* = *diag*(*σ*_1_^2^, …, *σ*_*q*_^2^). These parameters (e.g. variance components) and the fixed effects *β* are usually estimated by maximum likelihood (ML) or restricted maximum likelihood (REML) estimation [6]. The log-likelihood of LMM using all the data is

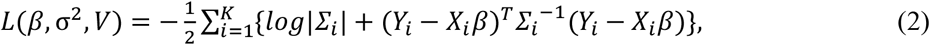

where *X*_*i*_ and *Y*_*i*_ are the covariate matrix and the outcome vector of the *i*^th^ site, |. | is the matrix determinant and 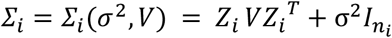.

The maximum likelihood estimation can be further simplified by profiling out *β* and σ^2^ from (2). Denote *Θ* = *V*/*σ*^2^, given *Θ*, the estimation of *β* and σ^2^ are

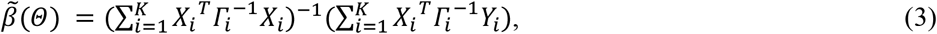

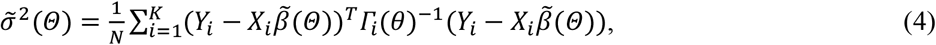

where 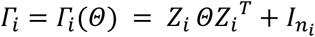. Thus the profile log-likelihood with respect to only *Θ* is

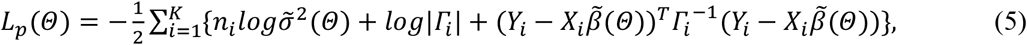

and the restricted profile log-likelihood is

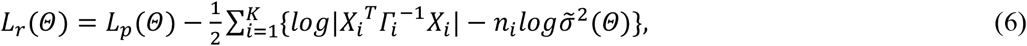

The ML or REML estimate of *Θ*can be obtained by maximizing (5) or (6). The estimates of *β* and σ^2^ can be subsequently obtained by (3) and (4). We denote these estimates as 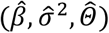. The variance of the estimated fixed effects 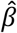 is thus

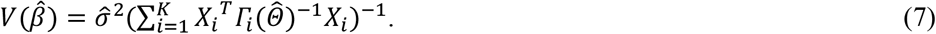

or the sandwich estimator:

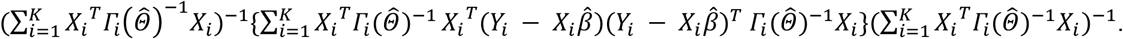

### 2.2 Distributed linear mixed model

It’s easy to see from (5) that there is no closed-form estimation for LMM. Thus unlike in the ordinary linear model [7], the LMM estimation is not trivial to be distributed to each site losslessly. Fortunately, with some linear algebra, we can disentangle the data (*Y*_*i*_, *X*_*i*_) and the parameters *Θ* in |*Γ*_*i*_| and *Γ*_*i*_^−1^and thus reconstruct the profile log-likelihood (5) without communicating individual patient data. Specifically, we utilize the Woodbury matrix identity [8] to obtain

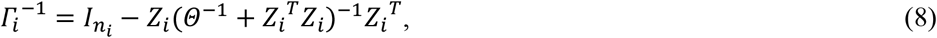

and the matrix determinant lemma [9] to obtain

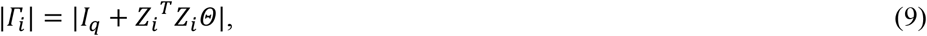

where *I*_*q*_ is the *q* × *q* identity matrix. We focus on the situation that the covariates in *Z* are a subset of that in *X*. The more general case is similar and will be elaborated in the Appendix. We require the *i*^th^ site to contribute some summary statistics, i.e. the *p* × *p* matrix *S*_*i*_ ^*X*^ = *X*_*i*_^*T*^*X*_*i*_, the *p* × 1 vector *S*_*i*_ ^*Xy*^ = *X*_*i*_ ^*T*^*y*_*i*_, the scalar *s*_*i*_^*Y*^ = *y*_*i*_^*T*^ *y*_*i*_ and the sample size *n*_*i*_. The exact log-likelihood (5) (or a restricted likelihood (6)) can then be reconstructed using these summary statistics. The details of the reconstruction are in the Appendix. The result by the DLMM algorithm is thus identical to that of the pooled LMM analysis, see Figure 2.

**Figure 2.**
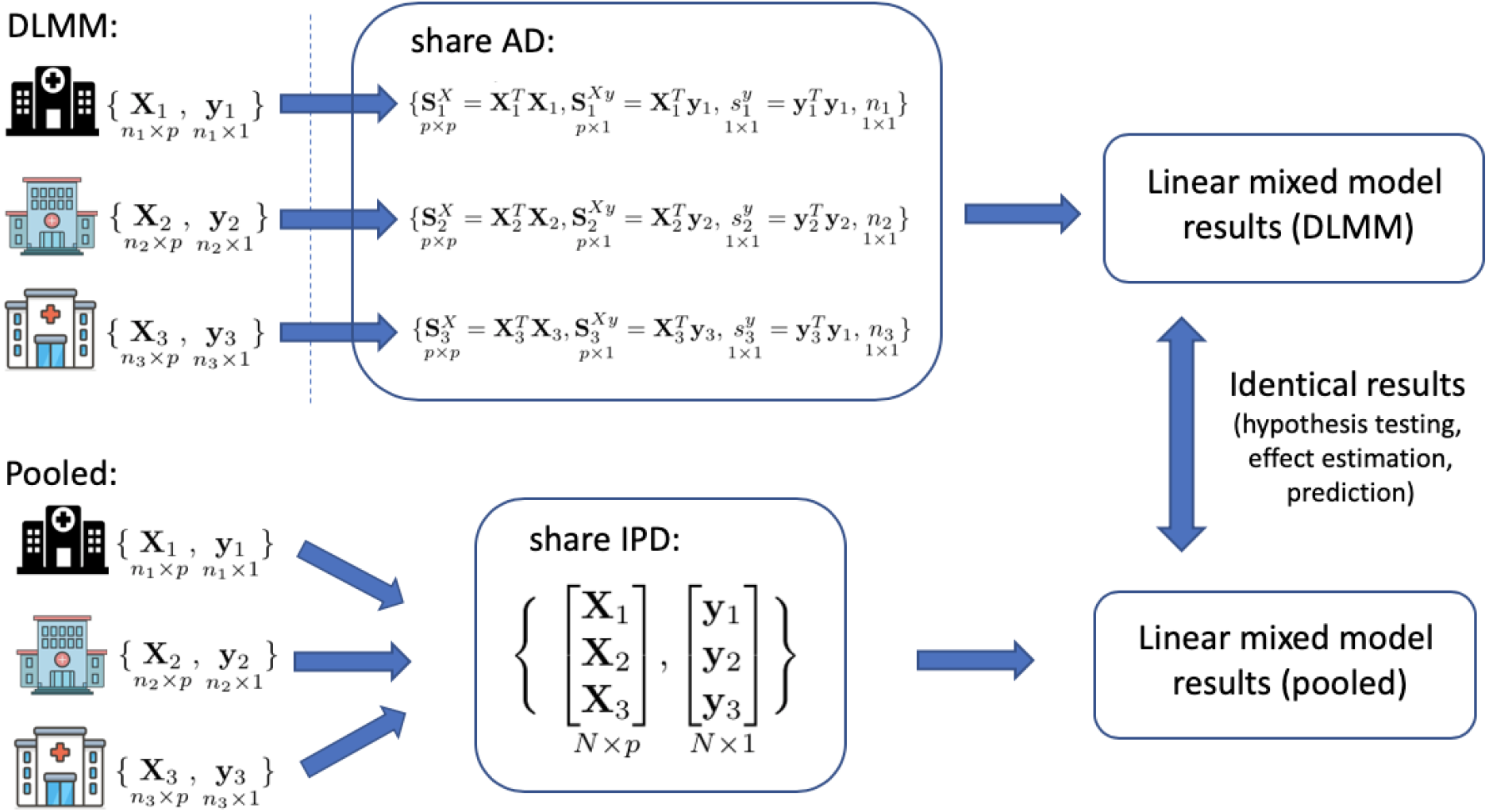
Schematic overview of the proposed algorithm for distributed linear mixed model (DLMM). The linear mixed model takes into account the heterogeneity of the effect of the covariates *X* on the continuous outcome *y* across sites. The proposed distributed algorithm achieves identical results as pooling the individual patient data (IPD) from all sites, by requiring only aggregated data (AD) *S*_*i*_^*X*^, *S*_*i*_^*Xy*^, *s*_*i*_^*y*^and sample size *n*_*i*_ from the *i*^th^ site. The distributed algorithm is privacy-preserving as only summary statistics (i.e. *p* × *p* matrices, *p* × 1 vectors and scalars) are being communicated.

### 2.3 Selection of variance components

We test the significance of random effects of each individual covariate by likelihood ratio test. For simplicity we assume the potential random effects are independent and the random intercept always exists, i.e. *V* = *diag*(*σ*_1_^2^, …, *σ*_*q*_^2^) and *σ*_1_^2^ > 0, for the covariate corresponding to variance component *σ*_*k*_^2^, *k* ≥ 2, we test

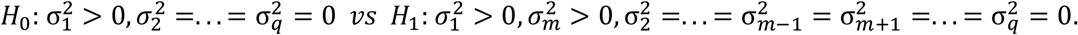

The likelihood ratio test (LRT) gives the likelihood ratio

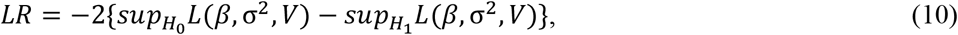

follows a 50:50 mixture of *χ*_0_^2^ and *χ*_1_^2^[10,11]. Notice both the log-likelihoods in (10) can be reconstructed by the communicated summary statistics.

### 2.4 Best linear unbiased predictors for the random effects

Finally, the BLUP [6] of the random effects *u*_*i*_ at the *i*^th^ site is

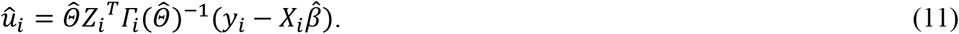

Conditioning on *X*_*i*_, +_*i*_has mean zero and covariance matrix

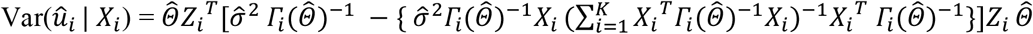

Since we are more interested in prediction of *u*_*i*_, it is more appropriate to use prediction intervals as below

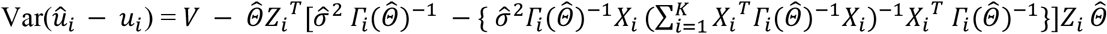

We summarize the analysis with the proposed DLMM algorithm as in Algorithm 1.

#### Algorithm 1. Analysis with the distributed linear mixed model algorithm

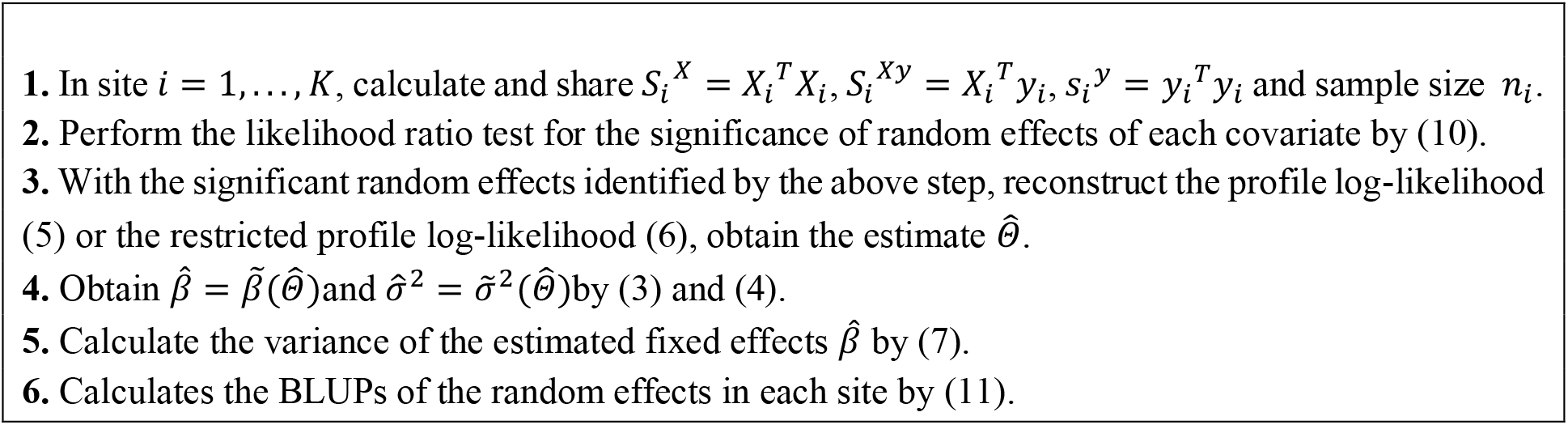

## 3. Multi-site analysis of COVID-19 hospitalization length of stay

We demonstrate the utility and lossless property of the DLMM method by studying the association of length of stay of COVID-19 hospitalization with patients’ demographic and clinical characteristics. We emphasize that this example is for illustrative purposes only and to this end we considered only covariates that have already been well-documented in the literature.

We identified patients who were admitted as inpatients to a hospital with a primary or secondary diagnosis of COVID-19 between January 1, 2020 and September 30, 2020. The data are collected from *K* = 538 sites in the UnitedHealth Group Clinical Research Database and the total number of patients is 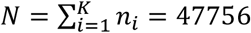. The detailed inclusion criteria is in Appendix Figure 1A. We treat length of stay as a continuous outcome. The demographic characteristics include age, gender and race and the clinical characteristics include a history of cancer, chronic obstructive pulmonary disease (COPD), heart disease, hypertension, hyperlipidemia, kidney disease and obesity. Charlson comorbidity index score is also included as a measure of the overall patient’s burden of diseases; the higher the score, the more severe the patient’s health condition is. We provide the details of the definition of the characteristics in the Appendix Table 2A.

**Figure 2.**
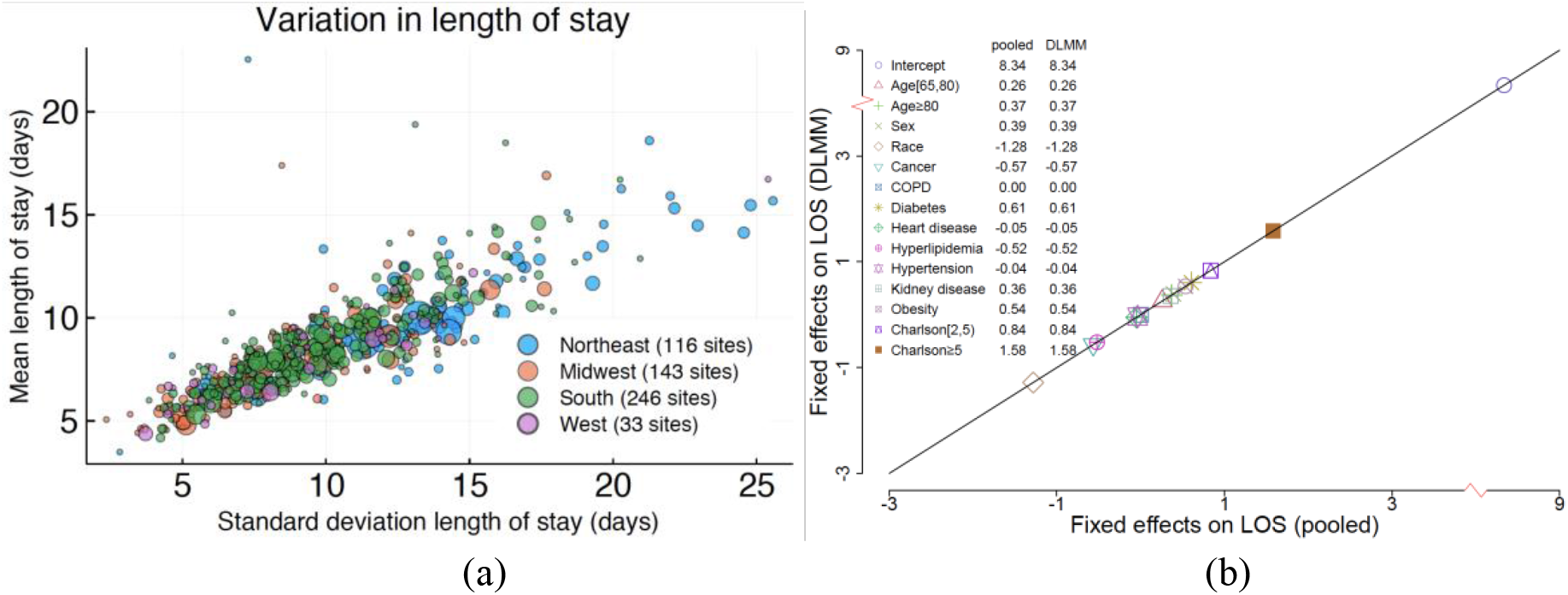
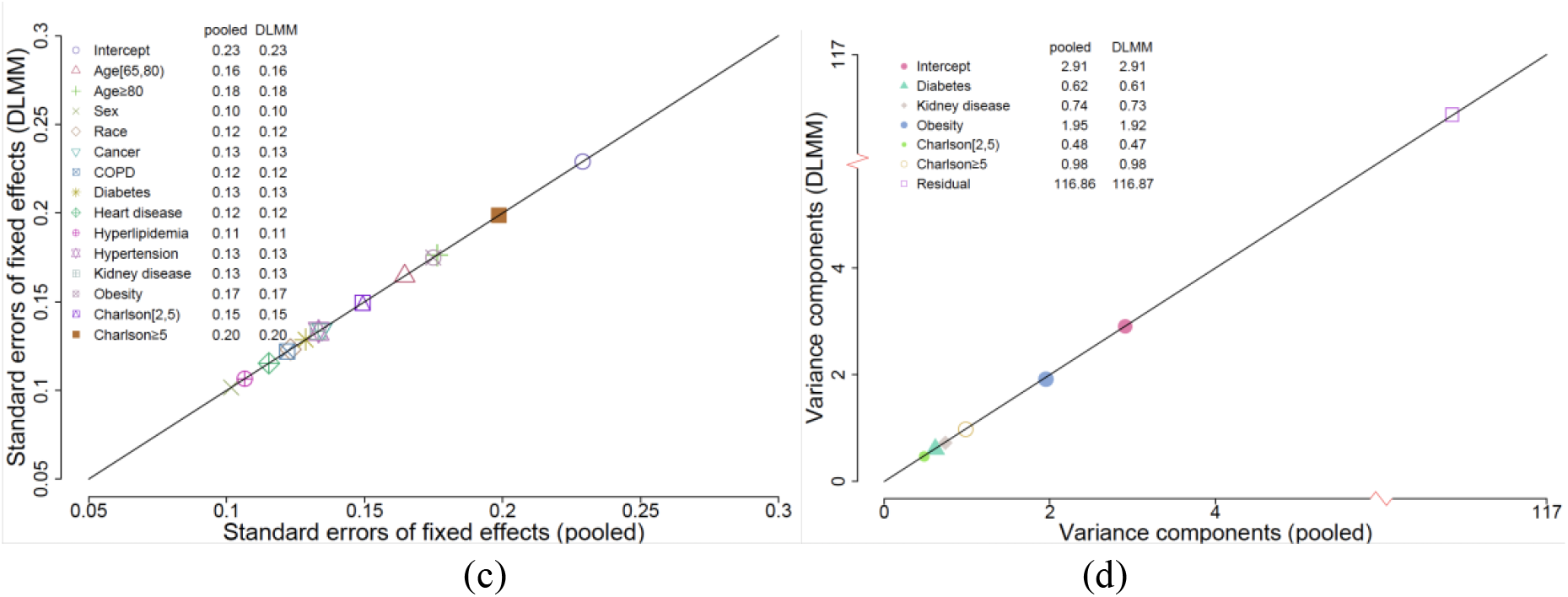
(a) The mean and standard deviation of length of stay of 47756 hospitalized COVID-19 patients from 538 hospitals. The data are collected from a single large US insurer and separated into their respective hospital sites to illustrate the algorithm. The area of each dot is proportional to the number of patients at that hospital and color represents the region. (b) Fixed effects estimation of linear mixed model by the proposed DLMM algorithm vs the pooled analysis. (c) Fixed effects’ standard error estimation of linear mixed model by the proposed DLMM algorithm vs the pooled analysis. (d) Variance components estimation of linear mixed model by the proposed DLMM algorithm vs the pooled analysis.

We select the covariate-specific random effects *u*_*im*_, *m* = 1, …, *q*, as described in Section 2.3. For simplicity we assume the random effect of different covariates are independent, i.e. *V* = *diag*(*σ*_1_^2^, …, *σ*_*q*_^2^). We select the covariates for which the corresponding p-value ≤0.05. In particular, we select random slopes for obesity, diabetes, kidney-diseases, and charlosn scores. A LMM with random intercept and random effects for obesity, diabetes, kidney-diseases, and charlosn scores is then fitted by either pooling the IPD together, or the proposed DLMM algorithm. We also calculate the BLUPs and the prediction intervals of the random effects at each site by (11) in Section 2.4.

We compare the result of the pooled analysis and the distributed algorithm in Figure 2 and Appendix Figure 3A. Specifically, the estimation of the fixed effects, their standard errors, the variance components are shown to be identical by the pooled analysis or the distributed algorithm. The estimated BLUPs by either the pooled analysis or the distributed algorithm are also shown to be identical in Figure 3A. The forest plot of fixed effects estimation and BLUPs of the random effects at a specific site are shown in Figure 3. Notice male, older age (≥80), higher Charlson score, obesity, prevalence of diabetes, and kidney diseases are shown to be significantly associated with longer COVID-19 hospitalization and Caucasian race, compared to others, is significantly associated with shorter COVID-19 hospitalization. The results match with that of literature for most of the covariates [16-19].

**Figure 3.**
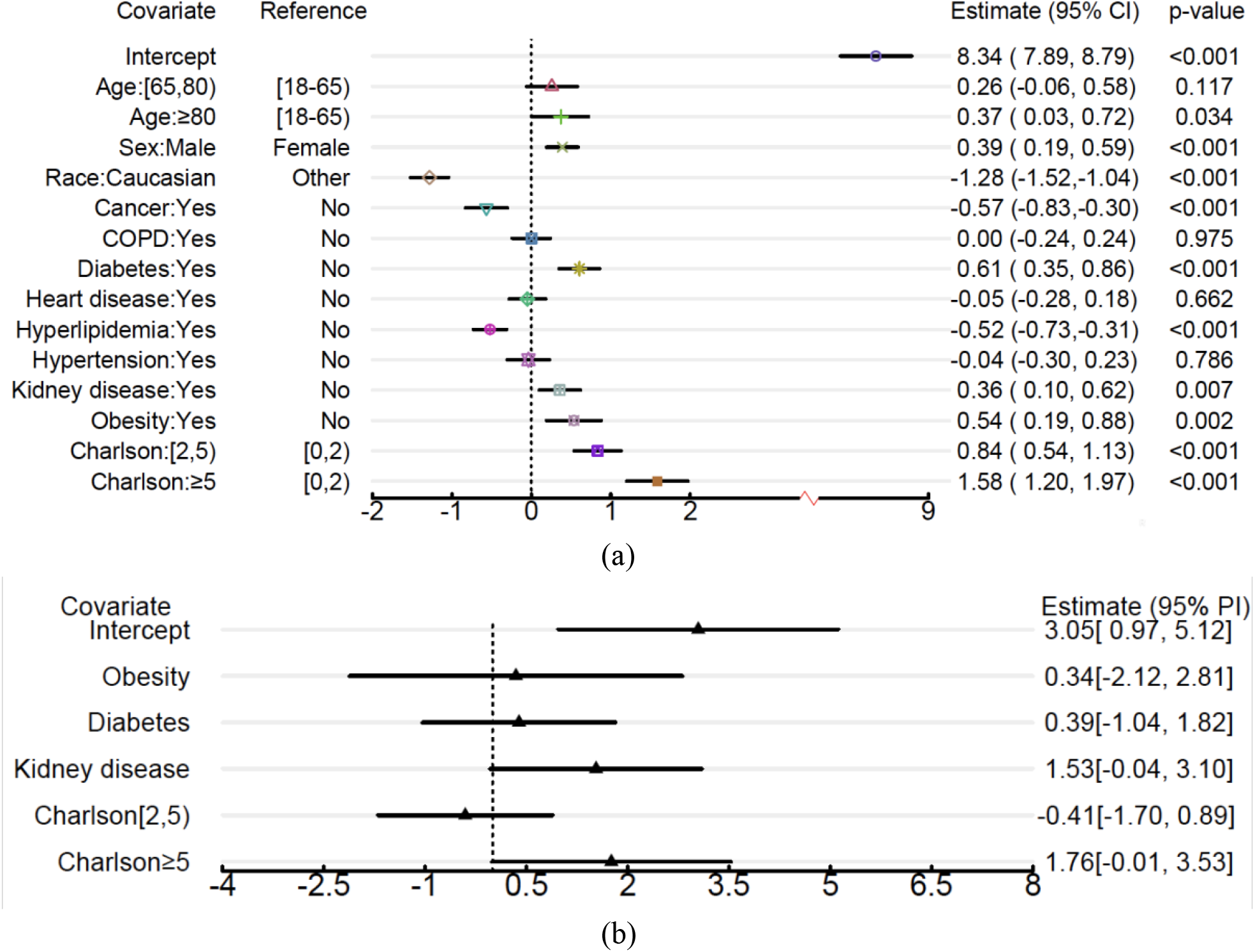
(a) Fixed effects of demographic and clinical characteristics on COVID-19 hospitalization length of stay. A vertical reference line is drawn for convenience in comparison. Reported are the estimated effect sizes, 95% confidence intervals and corresponding p-values based on Wald test. (b) BLUPs for random effects corresponding to a site located in the south region; reported are 95% prediction intervals.

## 4. Discussion and future work

Special care must be taken with healthcare data in order to preserve patient privacy. Anonymizing data while preserving features that are important for understanding an individual’s health is highly non-trivial. In addition, large, representative datasets are especially scarce. Distributed models solve the privacy issue by requiring that only summary level statistics are shared. The one-shot model presented here requires only the *p* × *p* matrix of summary statistics, sample size, and *p*-dimensional vector be sent once. This allows for efficient sharing to build models for various applications across healthcare where data may remain completely protected by eliminating the need for data pooling at a central source. By considering a large, more diverse sample from multiple sites we expect a more robust outcome which benefits all institutions.

## Data Availability

The data are proprietary and are not available for public use but can be made available under a data use agreement to confirm the findings of the current study.

## 5. Acknowledgement

## Appendix

### A1. Reconstruction of the (restricted) LMM likelihood

In the case that the random effects covariates in *Z* is not a subset of the fixed effects covariates in *X*, the proposed DLMM algorithm requires the *i*^*th*^ site to communicate

- *p* × *p matrix S*_*i*_^*X*^ = *X*_*i*_^*T*^ *X*_*i*_, *p* × *q matrix S*_*i*_^*XZ*^ = (*S*_*i*_^*ZX*^)^*T*^= *X*_*i*_^*T*^ *Z*_*i*_,
- *p* − *dim vector S*_*i*_^*Xy*^ = (*S*_*i*_^*yX*^)^*T*^ = *X*_*i*_^*T*^ *y*_*i*_, *q* − *dim vector S*_*i*_^*Zy*^ = (*S*_*i*_^*yz*^)^*T*^ = *Z*_*i*_^*T*^ *y*_*i*_,
- *scalar S*_*i*_^*y*^ = *y*_*i*_^*T*^ *y*_*i*_, *sample size n*_*i*_,

for reconstructing the (restricted) LMM likelihood.

Below are the details of the reconstruction. By (8),

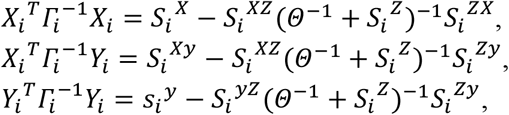

thus 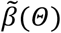 and 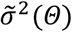 can be reconstructed by (3) and (4). Notice the unknown parameters are contained only in *Θ* and are separated from the summary statistics. Therefore, the profile log-likelihood (5) with respect to *Θ* can be reconstructed as

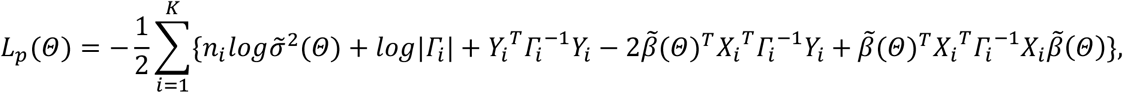

where |*Γ*_*i*_| = |*I*_*q*_ + *S*_*i*_ ^*Z*^*Θ*| according to (9). The restricted profile log-likelihood (6) can also be reconstructed in the same way.

### A2. Further information on data sources

#### A2.1 Standardization of data entry and data structure

Medical and pharmacy claims data are captured, predominantly electronically, from sites of care seeking third-party reimbursement for both Medicare and commercial plans using the industry standard data collection forms HCFA/CMS-1500 for facility claims, UB04/CMS-1450 for professional services and outpatient claims, and NCPDP for pharmacy claims or their electronic equivalents. Structured data from these standardized forms are coded using the International Classification of Diseases, Tenth Revision, Clinical Modification (ICD-10-CM), National Drug Codes (NDC), Current Procedural Terminology (CPT) codes, and Logical Observation Identifiers Names and Codes (LOINC) codes, and Diagnosis Related Groups (DRG). This nomenclature ensures consistency of data collection across geographic regions, health systems, and payers throughout the United States.

#### A2.2 Methods to Control for Errors in Sampling and Data Collection

Claims that do not adhere to the form or coding standards described above are rejected from reimbursement, minimizing the risk that inappropriately structured data are included in the database. Data specific to SARS-CoV-2 and COVID-19 has an additional Quality Control layer to control for errors in sampling and data collection; this is described below in the section on Quality Control.

**Figure 1A.**
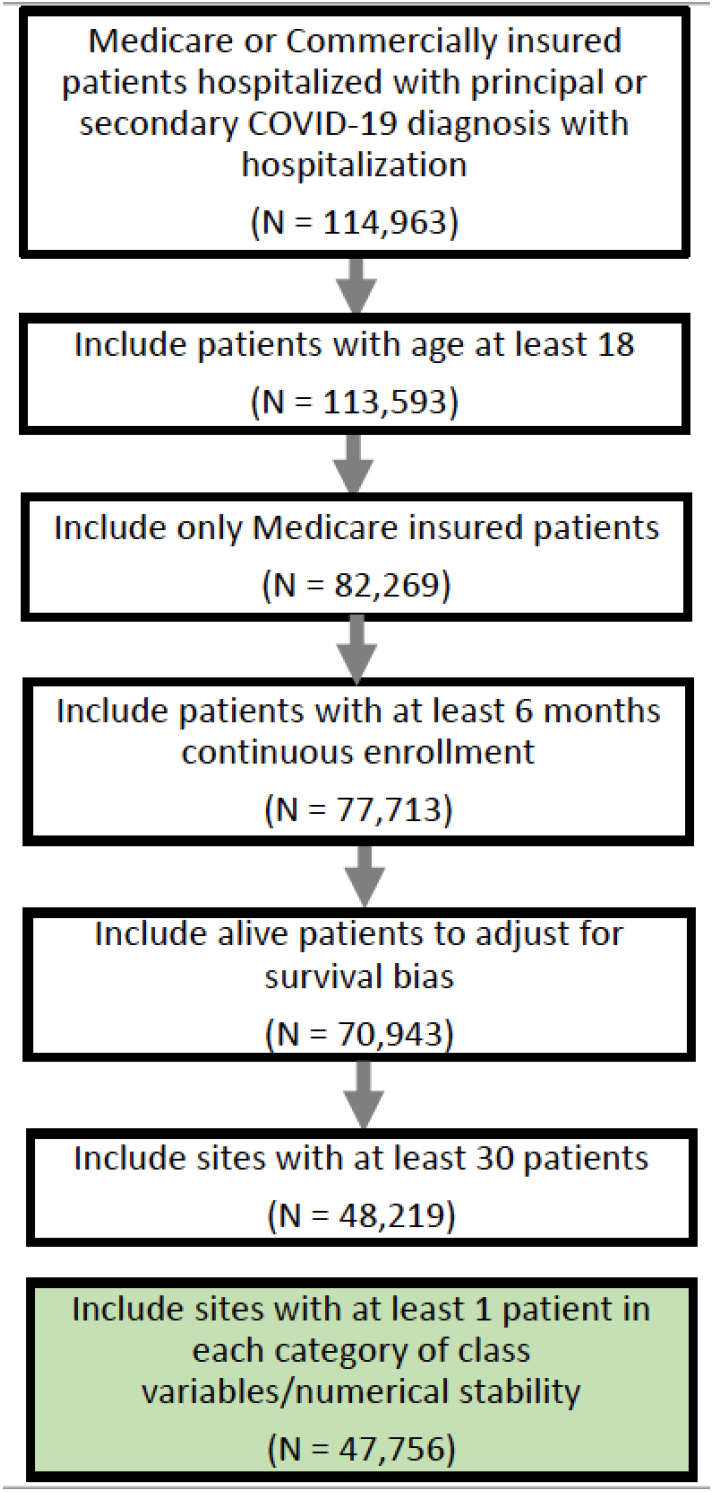
Flow chart of cohort definition for the COVID-19 hospitalization length of stay study using UHG claims data.

#### A2.3 Quality control

A COVID-19 data source-specific layer of quality control is also present, given the rapidly evolving situation. Members with a qualified COVID-19 related hospital admission are included in the report when any diagnosis matches qualified ICD-10 codes of U071, U072, or B9729. Suspected COVID-19 inpatient cases are manually reviewed daily by health plan clinical staff via clinical notes to determine an individual’s COVID-19 status. Each case is then manually flagged as either negative, confirmed, presumed positive, or needs clinical review. If a case is confirmed, it is not reviewed again. If a case is listed as negative or unknown, it is periodically reviewed for changes in the record. All others are reviewed and updated daily.

#### A2.4 Data Sharing

The data are proprietary and are not available for public use but, under certain conditions, may be made available to editors and their approved auditors under a data use agreement to confirm the findings of the current study.

**Figure 2A.**
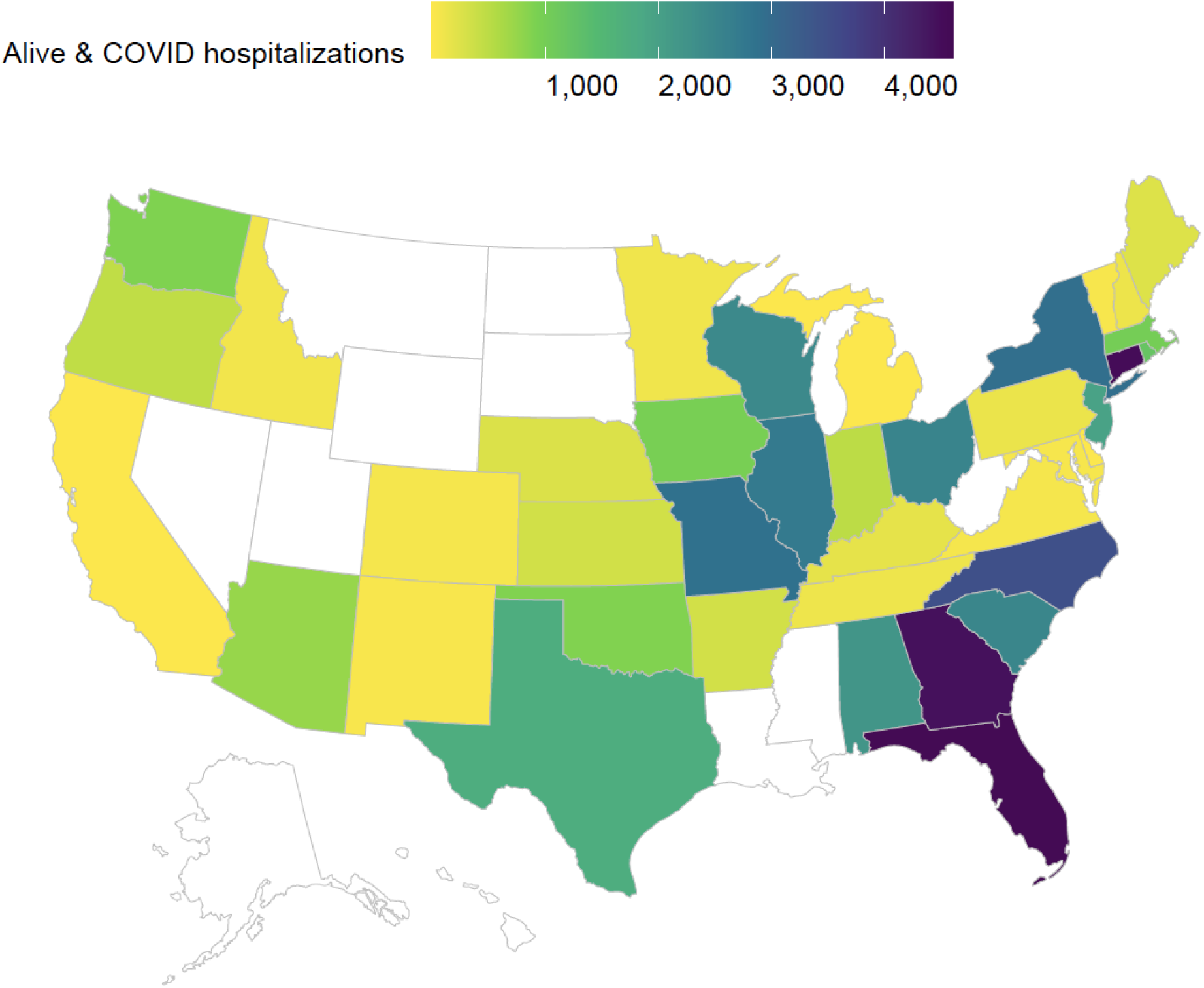
COVID-19 inpatient case distribution: number of hospitalizations by state. Data are extracted from UHG Clinical Research Database from Jan 1 to Sep 30, 2020.

**Table 1A.** UHG COVID-19 hospitalized patients characteristics.

|  |  |
| --- | --- |
| **Patients Number** | **47756** |
| **Hospitals by Region, count (%) Total** | 538 (100%) |
| Mid-west | 143 (26.6%) |
| North-east | 116 (21.6%) |
| South | 246 (45.7%) |
| West | 33 (6.13%) |
| **Patient Level Characteristics** |  |
| **Age category, count (%):**[18, 65) | 5638 (11.8%) |
| [65, 80) | 24571 (51.5%) |
| ≥ 85 | 17547 (36.7%) |
| **Gender, count (%):** Male (%) | 21625 (45.3%) |
| Female (%) | 26131 (54.7%) |
| **Race, count (%):** Caucasian (%) | 35083 (73.5%) |
| Other/unknown (%) | 12673 (26.5%) |
| **Charlson Score, count (%):**[0, 2) | 12578 (26.3%) |
| [2, 5) | 16805 (35.2%) |
| ≥ 5 | 18373 (38.5%) |
| **Comorbidities, count (%):**Cancer | 9612 (20.1%) |
| Chronic Obstructive Pulmonary Disease | 12035 (25.2%) |
| Diabetes | 21327 (44.7%) |
| Heart disease | 28586 (59.9%) |
| Hyperlipidemia | 28954 (60.6%) |
| Hypertension | 37986 (79.5%) |
| Kidney disease | 17664 (36.5%) |
| Obesity | 5865 (12.3%) |
| **Patient Outcome: Length of Stay in days, mean (sd)** | (8.6, 11.1) |

**Table 2A.** ICD-10-CM codes used to calculate Charlson comorbidity score.

| Comorbidity | ICD-10-CM Codes |
| --- | --- |
| Acquired immunodeficiency syndrome (AIDS) | B20, B21, B22, B24 |
| Arthritis | M05, M06, M315, M32, M33, M34, M351, M353, M360 |
| Cerebrovascular Disease | G45, G46, H340, I60, I61, I62, I63, I64, I65, I66, I67, I68, I69 |
| Congestive Heart Failure (CHF) | I099, I110, I130, I132, I255, I420, I425, I426, I427, I428, I429, I43, I50, P290 |
| Chronic obstructive pulmonary disease (COPD) | I278, I279, J40, J41, J42, J43, J44, J45, J46, J47, J60, J61, J62, J63, J64, J65, J66, J67, J684, J701, J703 |
| Dementia | F00, F01, F02, F03, F051, G30, G311 |
| Diabetes | E100, E101, E106, E108, E109, E110, E111, E116, E118, E119, E120, E121, E126, E128, E129, E130, E131, E136, E138, E139, E140, E141, E146, E148, E149 |
| Diabetes with complications | E102, E103, E104, E105, E107, E112, E113, E114, E115, E117, E122, E123, E124, E125, E127, E132, E133, E134, E135, E137, E142, E143, E144, E145, E147 |
| Mild Liver Disease | B18, K700, K701, K702, K703, K709, K713, K714, K715, K717, K73, K74, K760, K762, K763, K764, K768, K769, Z944 |
| Moderate/ Severe Liver Disease | I850, I859, I864, I982, K704, K711, K721, K729, K765, K766, K767 |
| Metastatic solid malignancy | C77, C78, C79, C80 |
| Myocardial infarction | I21, I22, I252 |
| Paralysis | G041, G114, G801, G802, G81, G82, G830, G831, G832, G833, G834, G839 |
| Peripheral Vascular Disease | I70, I71, I731, I738, I739, I771, I790, I792, K551, K558, K559, Z958, Z959 |
| Peptic Ulcer Disease | K25, K26, K27, K28 |
| Renal Disease | I120, I131, N032, N033, N034, N035, N036, N037, N052, N053, N054, N055, N056, N057, N18, N19, N250, Z490, Z491, Z492, Z940, Z992 |
| Tumor | C00, C01, C02, C03, C04, C05, C06, C07, C08, C09, C10, C11, C12, C13, C14, C15, C16, C17, C18, C19, C20, C21, C22, C23, C24, C25, C26, C30, C31, C32, C33, C34, C37, C38, C39, C40, C41, C43, C45, C46, C47, C48, C49, C50, C51, C52, C53, C54, C55, C56, C57, C58, C60, C61, C62, C63, C64, C65, C66, C67, C68, C69, C70, C71, C72, C73, C74, C75, C76, C81, C82, C83, C84, C85, C88, C90, C91, C92, C93, C94, C95, C96, C97 |

**Figure 3A.**
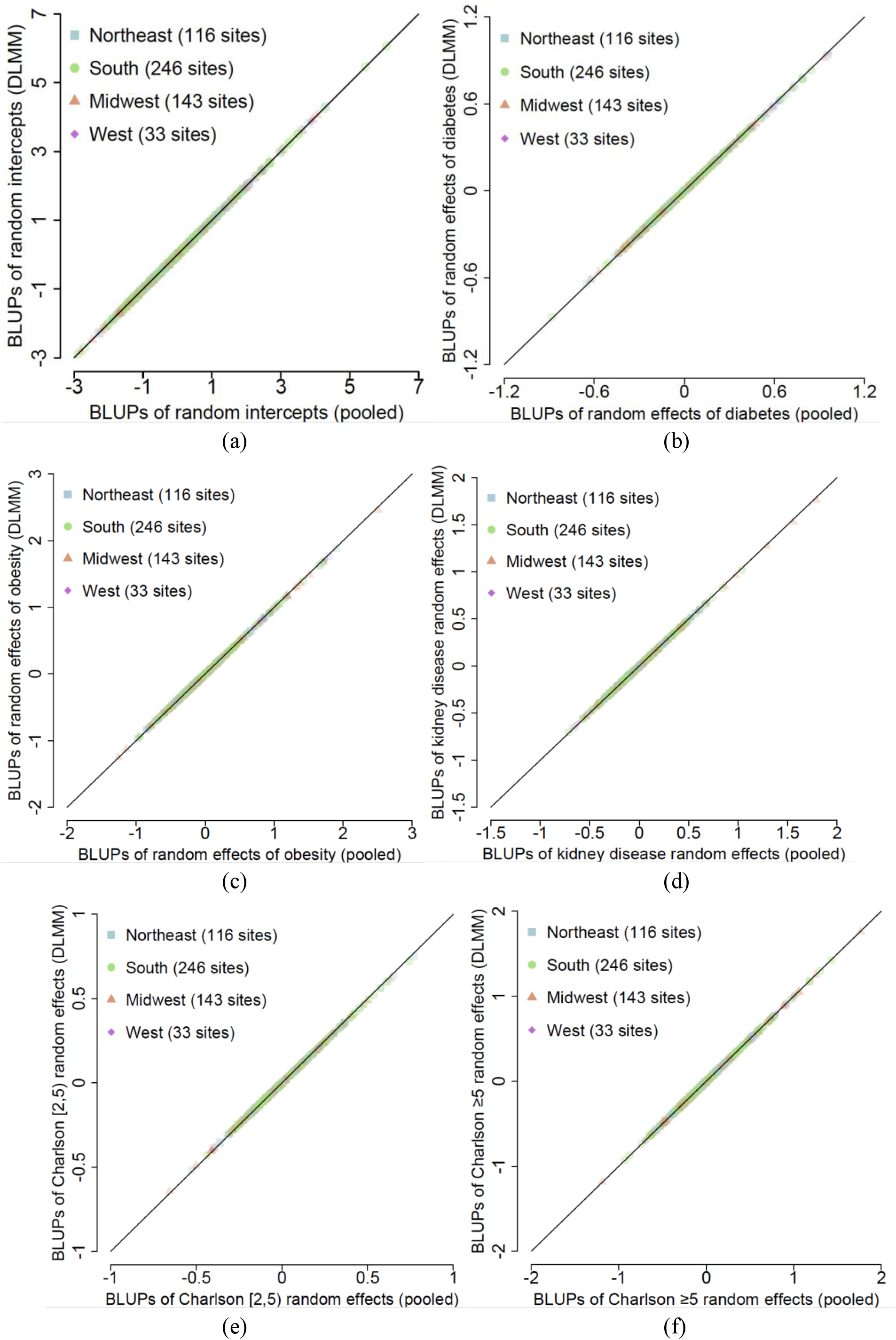
Comparison of the best linear unbiased predictors (BLUPs) of the random effects by pooled and DLMM methods. The BLUPs are obtained from a linear mixed model with COVID-19 hospitalization as the outcome and demographics and comorbidity variables as covariates. Diabetes, obesity, kidney disease and Charlson score are selected as having significant random effects.

## References

1. Kimmel SE, Califf RM, Dean NE, Goodman SN, Ogburn EL. COVID-19 Clinical Trials: A Teachable Moment for Improving Our Research Infrastructure and Relevance. Annals of Internal Medicine. 2020 Jun 16.

2. Becchetti, Leonardo and Conzo, Gianluigi and Conzo, Pierluigi and Salustri, Francesco, Understanding the Heterogeneity of Adverse COVID-19 Outcomes: The Role of Poor Quality of Air and Lockdown Decisions (April 10, 2020). Available at SSRN: http://dx.doi.org/10.2139/ssrn.3572548.

3. Chen Q, Allot A, Lu Z. Keep up with the latest coronavirus research. Nature. 2020 Mar;579(7798):193.

4. Ogburn EL, Bierer BE, Brookmeyer R, Choirat C, Dean NE, De Gruttola V, Ellenberg SS, Halloran ME, Hanley Jr DF, Lee JK, Wang R. Aggregating data from COVID-19 trials. Science (New York, NY). 2020 Jun 12;368(6496):1198–9.

5. Williamson EJ, Walker AJ, Bhaskaran K, Bacon S, Bates C, Morton CE, Curtis HJ, Mehrkar A, Evans D, Inglesby P, Cockburn J. OpenSAFELY: factors associated with COVID-19 death in 17 million patients. Nature. 2020 Jul 8:1–1.

6. Ruppert, D., Wand, M.P. and Carroll, R.J., 2003. Semiparametric regression (No. 12). Cambridge university press.

7. Chen, Y., Dong, G., Han, J., Pei, J., Wah, B.W. and Wang, J., 2006. Regression cubes with lossless compression and aggregation. IEEE Transactions on Knowledge and Data Engineering, 18(12), pp.1585–1599.

8. Sherman, J. and Morrison, W.J., 1950. Adjustment of an inverse matrix corresponding to a change in one element of a given matrix. The Annals of Mathematical Statistics, 21(1), pp.124–127.

9. Ding, J. and Zhou, A., 2007. Eigenvalues of rank-one updated matrices with some applications. Applied Mathematics Letters, 20(12), pp.1223–1226.

10. Self SG, Liang KY. Asymptotic properties of maximum likelihood estimators and likelihood ratio tests under nonstandard conditions. Journal of the American Statistical Association. 1987 Jun 1;82(398):605–10.

11. Stram DO, Lee JW. Variance components testing in the longitudinal mixed effects model. Biometrics. 1994 Dec 1:1171–7.

12. Duan R, Boland MR, Liu Z, Liu Y, Chang HH, Xu H, Chu H, Schmid CH, Forrest CB, Holmes JH, Schuemie MJ. Learning from electronic health records across multiple sites: A communication-efficient and privacy-preserving distributed algorithm. Journal of the American Medical Informatics Association. 2020 Mar;27(3):376–85.

13. Duan, R., Luo, C., Schuemie, M. H., Tong, J., Liang, J. C., Chang, H. H., Boland, M. R., Bian, J., Xu, H., Holmes, J. H.. Learning from local to global-an efficient distributed algorithm for modeling time-to-event data. Journal of the American Medical Informatics Association. 2020 July; 27(7):1028–1036.

14. Lu, C.-L., Wang, S., Ji, Z., Wu, Y., Xiong, L., Jiang, X., and Ohno-Machado, L.. Webdisco: a web service for distributed cox model learning without patient-level data sharing. Journal of the American Medical Informatics Association 22, 1212–1219.

15. Wu, Y., Jiang, X., Kim, J., and Ohno-Machado, L.. G rid binary lo gistic re gression (glore): building shared models without sharing data. Journal of the American Medical Informatics Association 19, 758–764.

16. Sanyaolu A, Okorie C, Marinkovic A, Patidar R, Younis K, Desai P, Hosein Z, Padda I, Mangat J, Altaf M. Comorbidity and its Impact on Patients with COVID-19. Sn Comprehensive Clinical Medicine. 2020 Jun 25:1–8.

17. Price-Haywood EG, Burton J, Fort D, Seoane L. Hospitalization and mortality among black patients and white patients with Covid-19. New England Journal of Medicine. 2020 May 27.

18. Rees EM, Nightingale ES, Jafari Y, Waterlow NR, Clifford S, Pearson CA, Jombart T, Procter SR, Knight GM, CMMID Working Group. COVID-19 length of hospital stay: a systematic review and data synthesis. medRxiv preprint https://doi.org/10.1101/2020.04.30.20084780.

19. Wang L, He W, Yu X, Hu D, Bao M, Liu H, Zhou J, Jiang H. Coronavirus disease 2019 in elderly patients: Characteristics and prognostic factors based on 4-week follow-up. Journal of Infection. 2020 Mar 30.

